# High SARS-CoV-2 seroprevalence in rural Peru, 2021; a cross-sectional population-based study

**DOI:** 10.1101/2021.10.19.21265219

**Authors:** Andres Moreira-Soto, Johanna Maribel Pachamora Diaz, Lilian González-Auza, Xiomara Jeanleny Merino Merino, Alvaro Schwalb, Christian Drosten, Eduardo Gotuzzo, Michael Talledo, Heriberto Arévalo Ramirez, Roxana Peralta Delgado, Spassky Bocanegra Vargas, Jan Felix Drexler

## Abstract

Latin America has been severely affected by the COVID-19 pandemic. The COVID-19 burden in rural settings in Latin America is unclear. We performed a cross-sectional, population-based, random-selection SARS-CoV-2 serological study during March 2021 in the rural population of San Martin region, northern Peru. The study enrolled 563 persons from 288 houses across 10 provinces, reaching 0.19% of the total rural population of San Martin. Screening for SARS-CoV-2 IgG antibodies was done using a chemiluminescence immunoassay (CLIA) and reactive sera were confirmed using a SARS-CoV-2 surrogate virus neutralization test (sVNT). Validation using pre-pandemic sera from two regions of Peru showed false-positive results in the CLIA (23/84 sera; 27%), but not in the sVNT, highlighting the pitfalls of SARS-CoV-2 antibody testing in tropical regions and the high specificity of the two-step testing algorithm. An overall 59.0% seroprevalence (95% CI: 55-63%) corroborated intense SARS-CoV-2 spread in San Martin. Seroprevalence rates between the 10 provinces varied from 41.3-74.0% (95% CI: 30-84). Higher seroprevalence was neither associated with population size, population density, surface area, mean altitude or poverty index in spearman correlations. Seroprevalence and reported incidence diverged substantially between provinces, suggesting regional biases of COVID-19 surveillance data. Potentially, limited healthcare access due to environmental, geographic, economic, and cultural factors, might lead to undetected infections in rural populations. Additionally, test avoidance to evade mandatory quarantine might affect rural regions more than urban regions. Serologic diagnostics should be pursued in resource-limited settings to inform country-level surveillance, vaccination strategies and support control measures for COVID-19.

**Importance:** Latin America is a global hotspot of the COVID-19 pandemic. Serological studies in Latin America have been mostly performed in urban settings. Rural populations comprise 20% of the total Latin American population. Nevertheless, information of COVID-19 spread and transmission in rural settings is scarce. Using a representative population-based seroprevalence study, we detected a high seroprevalence in rural populations in San Martin, northern Peru in 2021, reaching 41 to 74 %. However, seroprevalence and reported incidence diverged substantially between regions, suggesting either limited healthcare access or test avoidance due to mandatory quarantine. Our results suggest that rural populations are highly affected by SARS-CoV-2 even though they are socio-demographically distinct from urban populations, and that highly specific serological diagnostics should be performed in resource-limited settings to support public-health strategies of COVID-19 surveillance and control.

## Background

Peru has been severely affected by the COVID-19 pandemic, with the highest mortality per capita reported worldwide since the start of the pandemic, reaching 6,132 deaths per million as of October 2021 (1, 2). The determinants of SARS-CoV-2 spread in Latin America are poorly defined. An epidemiological study from European and Asian urban centers correlated population density with increased SARS-CoV-2 infection rates (3). In contrast, epidemiological studies from Latin American urban centers yielded diverse prevalence estimates that seemed uncorrelated to population density, exemplified by three available seroprevalence studies from Peru, conducted during mid-late 2020. The first study from the Peruvian capital Lima (population: 9.5 million; density: 12,000/km^2^) reported a 21% seroprevalence rate (4). The second study from the Lambayeque department in northern Peru (population: 1.2 million; density: 84/km^2^) reported a 29% seroprevalence rate (5). The third study from Iquitos city, Peruvian Amazon (population: 470,000; density: 417/km^2^) reported a 70% seroprevalence rate, implying local herd immunity had been reached (6). Despite the high seroprevalence already during 2020, a second SARS-CoV-2 wave occurred in Iquitos in January 2021. Potentially, mobile susceptible populations in the Amazon region might explain occurrence of a second wave (6). Hypothetically, susceptible populations might correspond to rural and indigenous populations, comprising ≤19% of the total Latin American population based on World Bank estimates (https://data.worldbank.org/indicator/SP.RUR.TOTL.ZS?locations=ZJ). The high geographical dispersion and low population density of these communities, in addition to regular travelling to urban centers for commerce, might contribute to a differential dispersion of SARS-CoV-2 compared to urban settings.

### Seroprevalence study in San Martin

Our study focused on the rural region of San Martin, Peru (total population: 899,648 density: 18/km^2^). The San Martin department (51,000 km^2^) is located in the north of Peru (**Figure 1A**). The department encompasses diverse ecosystems such as valleys, Andes mountains and Amazon rainforests, reaching altitudes from 190 to 3,080 m. (**Figure 1A**) and comprises ≤5% of Peruvian forest cover. The San Martin population is divided into Mestizo (83.6%) followed by Quechua (5.3%) and Afro-Peruvian (4.9%) ethnicities as per census data of the National Institute of Statistics and Census of Peru (INEI; www.inei.gob.pe/).

**Figure 1.**
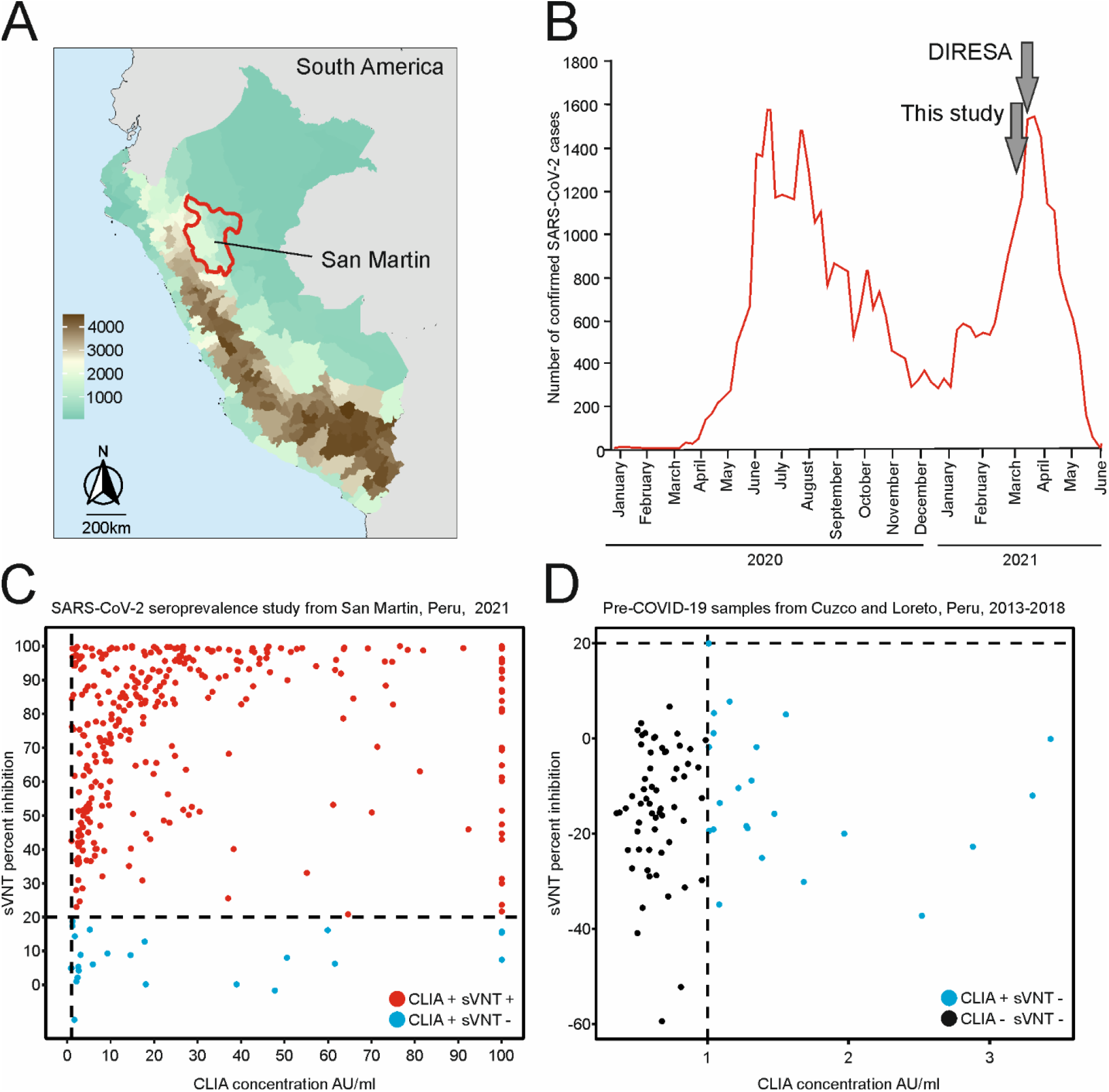
Epidemiological surveillance and serological diagnostics of SARS-CoV-2 from San Martin, Peru. (A) Mean altitude map of Peru. In red the department of San Martin. (B) Number of confirmed cases in San Martin as of June 2021. The time where incidence data and samples for the serological study were taken is marked with arrows. Surveillance data were gathered from https://diressanmartin.gob.pe/ and https://www.gob.pe/minsa/. Reactivity of serum samples from (C) SARS-CoV-2 seroprevalence study in San Martin in 2021 and (D) pre-COVID-19 cohort from 2013-2018 from Peru in a chemiluminescence immunoassay (CLIA) and a SARS-CoV-2 surrogate virus neutralization test (sVNT, GenScript, USA).

We performed a cross-sectional population-based random-selection study during March 2021 (institutional ethics committee Via Libre; number 6532-2021a) in the ten provinces of San Martin (**Figure 1B**). Sample size calculation considered the rural population of San Martin (286 988), a 95% confidence level and an estimated SARS-CoV-2 prevalence of 50%, reaching 576 individuals. As the average household in Peru consists of 4 inhabitants (www.inei.gob.pe/), the study aimed at sampling two individuals from 288 individual houses. First, a conglomerate of 100 rural houses was selected based on estimates of the census data of 2017 and on geographic information systems of the INEI. Next, a random household selection in the conglomerate was performed. Exclusion criteria encompassed population residing in collective dwellings such as barracks, police stations, convents, boarding schools and hotels, used as temporary housing of unrelated people with unstable population dynamics, age ≤5 years, skin lesions in the venous puncture site, usage of alcohol or psychoactive drugs, not being permanent resident and not signing the informed consent. Participation was voluntary and participant selection in households was performed using a kish grid (7). A 10 mL blood sample was taken after the informed consent was signed by the person or the caretaker if the person had ≤18 years of age. We obtained samples from 563 persons visiting 288 houses in the 10 provinces, comprising 0.19% of the total rural population of San Martin. The sampled cohort included persons aged 6-89 years (mean: 35.8, standard deviation (SD): 21.15) consistent with San Martin’s age distribution data from the INEI 2017 census (**Annex Figure 1;** www.inei.gob.pe/). From the cohort, 37.7% (212/563) were male and 62.3% (351/563) female, contrasting with the sex distribution in San Martin (51.3% male, 48.7% female). The difference between the distribution can be explained by the sampling strategy, limited at visiting the homes of the participants during the day due to safety and operational reasons, while males were mostly out working.

### Two-step serological testing and validation

Using a chemiluminescence immunoassay (CLIA) (SARS-CoV-2 S-RBD IgG kit, Snibe Diagnostic, China), we detected SARS-CoV-2-specific IgG antibodies in 63.6% of the samples (358/563) (**Figure 1C** and **Annex Figure 2)**. To validate the CLIA results, we used 84 pre-COVID-19 samples from our previous study in arbovirus serology in Peru (8) (**Figure 1D** and **Annex Figure 2**). The samples were collected in the municipalities of Cusco (altitude 3,400 m.) and Loreto (altitude 150 m.) during 2013 and 2018, respectively. A total of 27% (23/84; 11 from Cuzco and 12 from Loreto) of pre-COVID-19 samples yielded positive results in the CLIA screening test irrespective of geographical location (**Figure 1D** and **Annex Figure 2**). The apparently positive pre-COVID-19 samples showed statistically significantly lower signal concentrations in the CLIA than those samples from 2021 (median pre-COVID-19= 1.6 vs COVID-19= 32.81; t-test: p< 0.001) (**Annex Figure 2**), suggesting unspecific reactivity. Unspecific reactivity may be elicited by common-cold coronaviruses (9) or endemic tropical diseases such as Malaria, Dengue and Zika (10, 11). Therefore, a confirmatory SARS-CoV-2 surrogate virus neutralization test (sVNT, GenScript, USA) was performed in all CLIA-reactive samples. A total of 92.7% (332/358) of the 2021 CLIA-positive samples were confirmed using the sVNT which can be attributed to differential sensitivity of the tests. (**Figure 1C** and **Annex Figure 2**). None of the CLIA-positive pre-pandemic samples yielded positive results in the sVNT. These results corroborate that unspecific reactivity of serological tests in tropical areas must be carefully evaluated (10) and highlight robustness of our serological testing algorithm. Therefore, only samples yielding positive results in both serological assays were considered for further analyses in our study to guarantee specificity.

**Figure 2.**
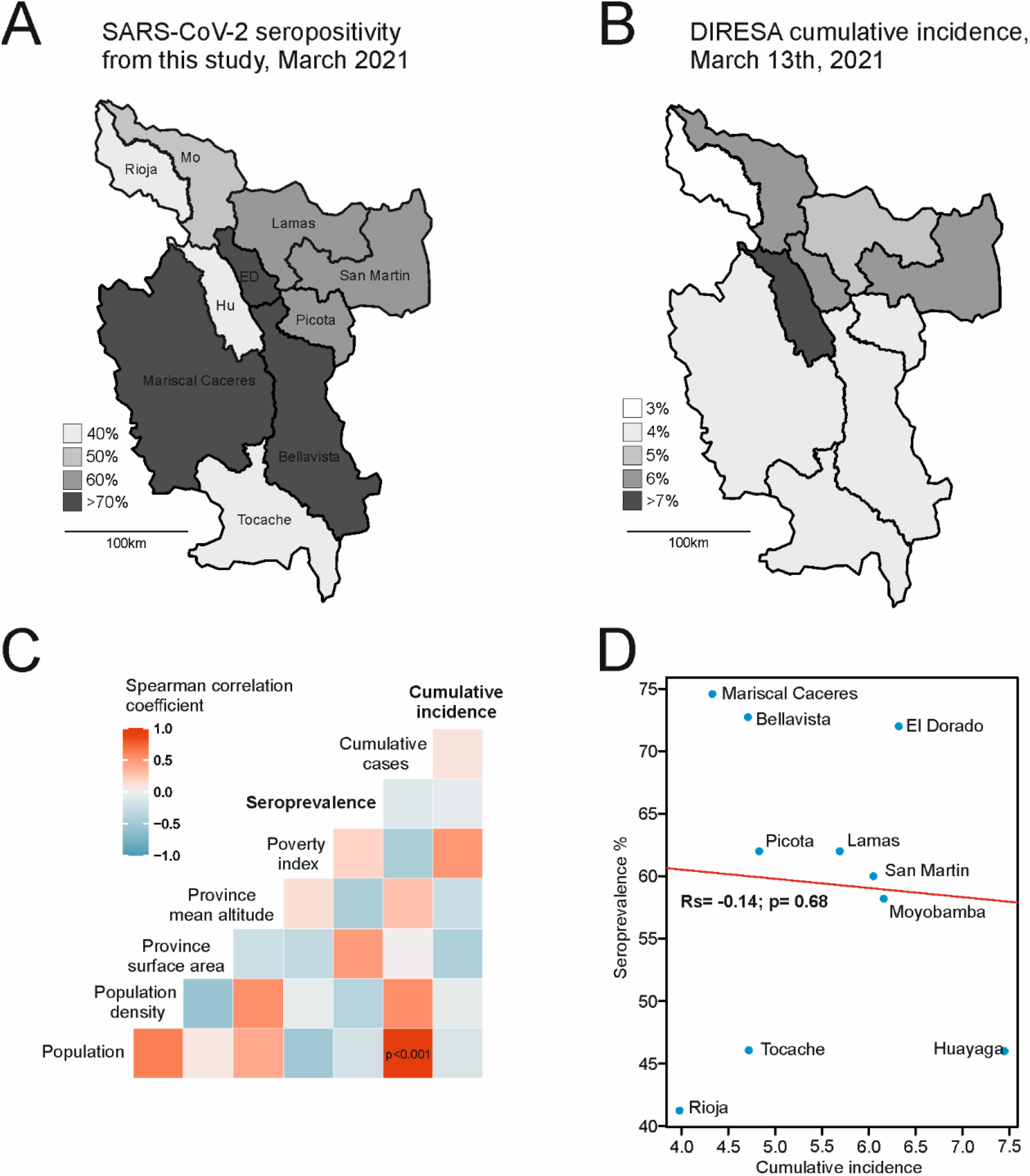
Correlation of seroprevalence and incidence data with different indicators. Comparison of serological (A) and incidence (B) data from San Martin. Seropositivity by provinces of San Martin. Hu= Huallaga, Mo= Moyobamba, ED= El Dorado. Right: confirmed cases by province from the Regional Health Directorate of Peru (DIRESA) as of 13/03/2021. Scale denotes percentage of cumulatively confirmed cases per total of the province population. (C) Heatmap of Spearman’s rank correlation test against different social, economic, and geographical indicators. Significant correlations are depicted inside the square. (D) Spearman’s rank correlation test of seroprevalence and cumulative incidence per province. Rs=Spearman’s correlation coefficient.

### Seroprevalence and statistical analyses

Overall, the seroprevalence for San Martin was 59.0% (332/563; 95% Confidence Interval (CI): 55-63%). No statistically significant difference of seroprevalence per sex was observed, using a chi-square test (*χ2*=0.05; p=0.83). However, a statistically significant difference was observed between the age of SARS-CoV-2-seropositive versus SARS-CoV-2-seronegative persons using a t-test (33 years (range: 6-89); versus 38 years (range: 7-57; p=0.01)). This observation was in concordance to the Iquitos study, likely associated with higher contact rates in younger age groups (6).

Seroprevalence between the 10 provinces varied from 41.3-74.0% (CI: 30-84%) (**Supplementary Table 1, Figure 2A** and **2B**). Higher seroprevalences were neither associated with population size (Spearman correlation test: r_s_=-0.26; p=0.46), nor population density (r_s_=-0.14; p=0.68), nor surface area (r_s_=0.33; p=0.34), nor mean altitude of the province (r_s_=-0.59; p=0.06), nor poverty index (r_s_=0.10; p=0.76) (**Figure 2C** and **Annex Figure 3**). Interestingly, the cumulative epidemiological surveillance data from the Regional Health Directorate of Peru (DIRESA) relying on notified cases of acute SARS-CoV-2 infection confirmed either by RT-PCR or an antigen test (**Figure 2B**), showed no correlation with our serological data although cumulatively measuring similar time spans, encompassing early 2020-March 2021 (r_s_=-0.14; p=0.68) (**Figure 1B** and **Figure 2D**). On the one hand, overall higher seroprevalence than reported cases is consistent with high numbers of undetected asymptomatic COVID-19 cases (12). On the other hand, we detected two provinces with a relatively higher divergence of seroprevalence compared to incidence data (Mariscal Caceres and Bellavista, **Figure 2A, 2B** and **2D**). Hypothetically, individuals in those provinces may lack healthcare access due to environmental, geographic, economic, and cultural factors (13), that might lead to more undetected infections in rural populations. First, test refusal to avoid mandatory quarantine or stigmatization has been reported for SARS-CoV-2 from different countries (14, 15). Test avoidance might be greater in rural regions from resource-limited areas, were a high share of the population work in jobs that do not allow quarantine for long periods of time and were lower incomes may force them to work and not visit or afford the hospital. Second, local health centers might have difficult access, or not be sufficiently equipped for the high COVID-19 burden (16), limiting access to diagnostics in rural areas. High infection rates in rural populations might be explained by economic, or cultural factors. Lack of access to basic housing elements such as sewage disposal system, as observed in rural populations in Ecuador may contribute to the high seroprevalence in rural Peru (17). A survey analyzing rural residents in the USA found that they were less likely to participate in COVID-19 preventive measures such as the use of masks in public or working from home, increasing the possibility of infection (18). Another province (Huallaga, **Figure 2A, 2B** and **2D**) showed higher cumulative incidence than other provinces for unknown reasons, substantiating inconsistencies between serological and incidence data.

Irrespective of the underlying reasons leading to the overall high seroprevalence detected, our data suggest a large number of undiagnosed COVID-19 cases potentially challenging test-trace-isolate interventions in the region (19). Previous studies in the USA have stressed that rural regions are particularly vulnerable to COVID-19, leading to higher mortality rates in rural than in urban regions, and with higher mortality rates associated with black and Hispanic populations (20, 21), suggesting further studies are needed in Latin American vulnerable populations. Limitations of our study include lack of metadata and potential sampling biases. However, thorough study design, exhaustive serological testing and high seroprevalence consistent with data from other Latin American settings suggest robustness of our results (6, 17). Serological diagnostics should be pursued in resource-limited settings to inform country-level surveillance, vaccination strategies and support control measures for COVID-19.

## Data Availability

All data produced in the present work are contained in the manuscript

## Acknowledgments

We thank the DIRESA workers that aided in the sample collection and all study participants. We thank Victor Carvalho for technical support.

## Supplementary Material

**Annex Table 1.**
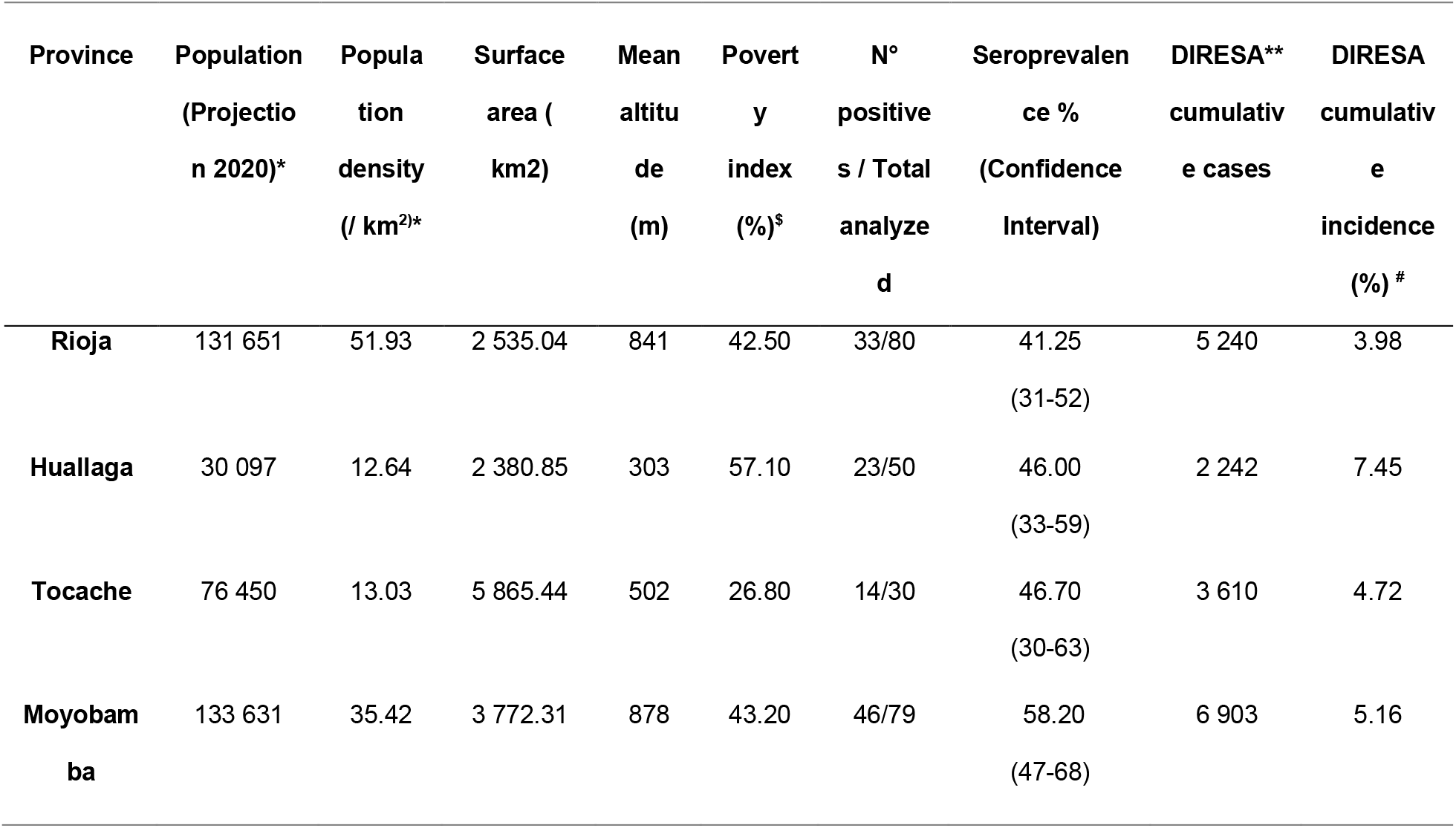
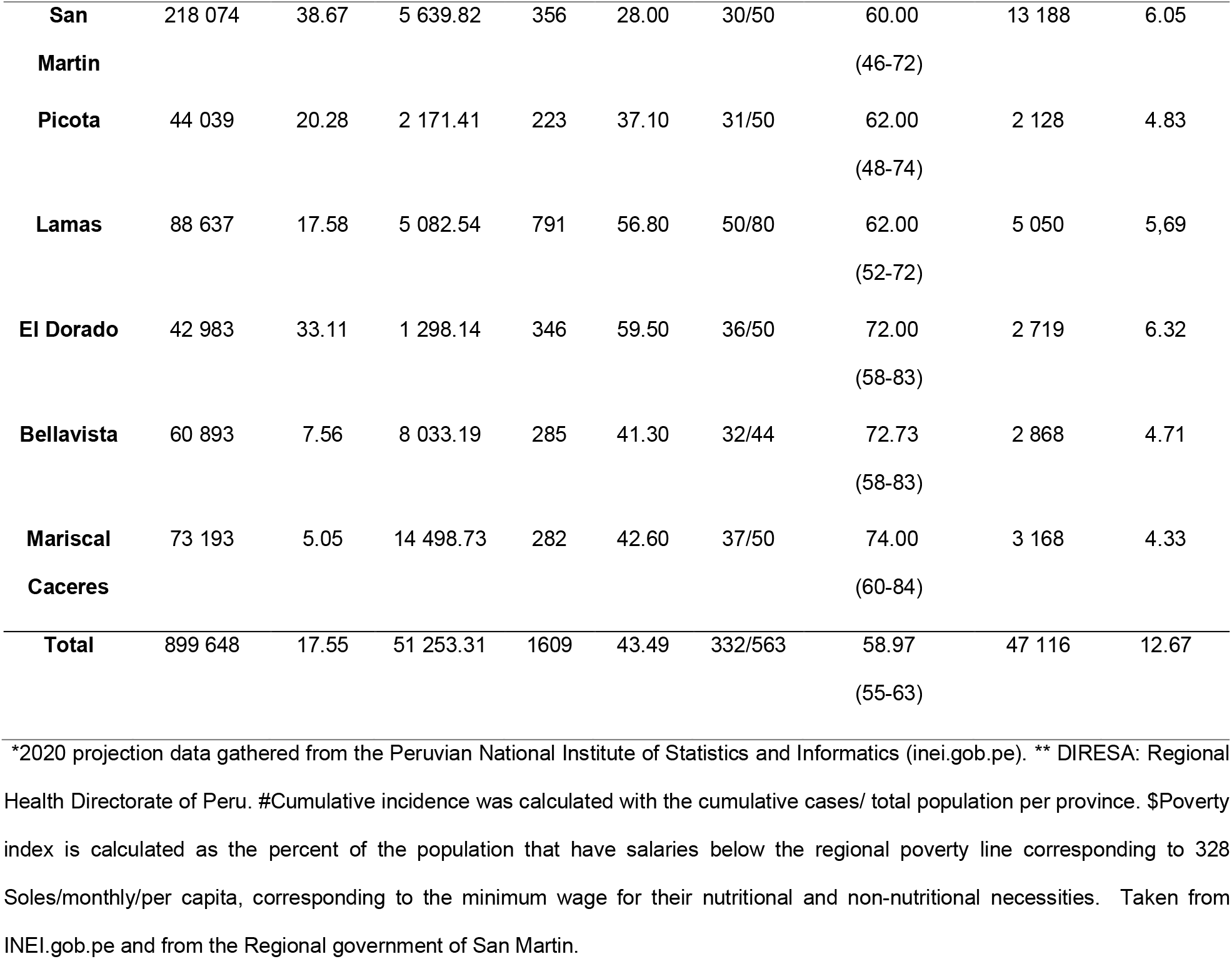
Demographics per province, seropositivity and incidence data from San Martin, Peru, 2021

**Annex Figure 1.**
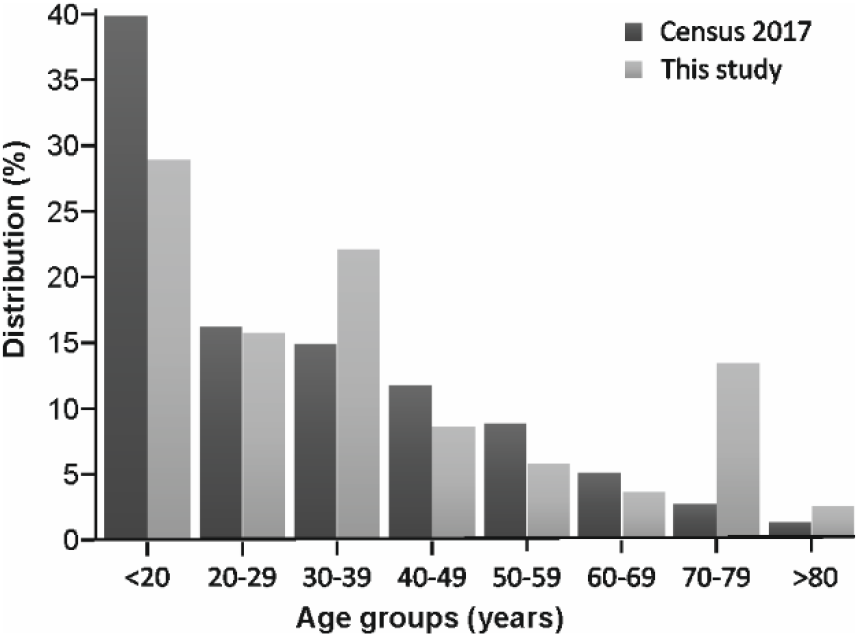
Age distribution from Census data in San Martin in comparison to this study. Data from the National Institute of Statistics and Informatics of Peru (https://www.inei.gob.pe/).

**Annex Figure 2.**
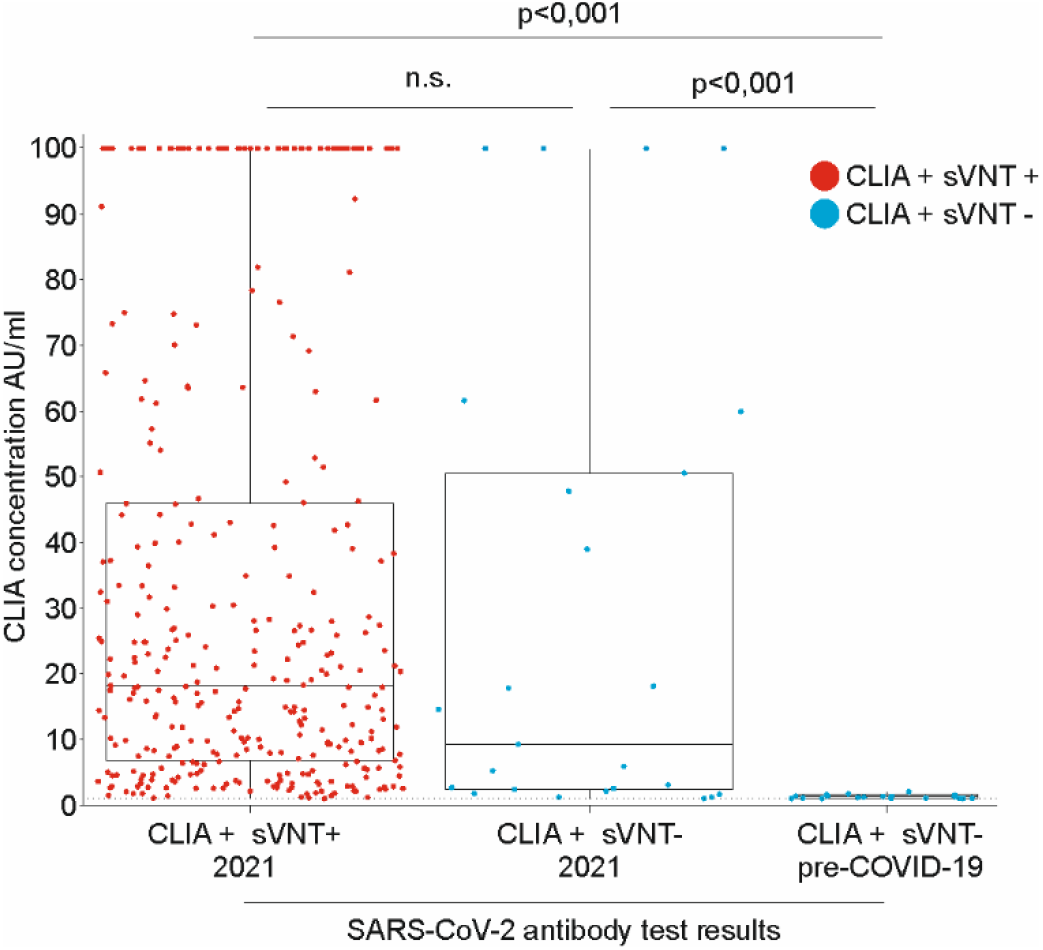
Validation of the testing algorithm using pre-COVID-19 samples from Peru. CLIA concentration comparison between SARS-CoV-2 positive samples from the 2021 cohort from this study and 84 pre-COVID-19 samples from Peru. Line inside the boxplots denotes the median. N.s. not significant.

**Annex Figure 3.**
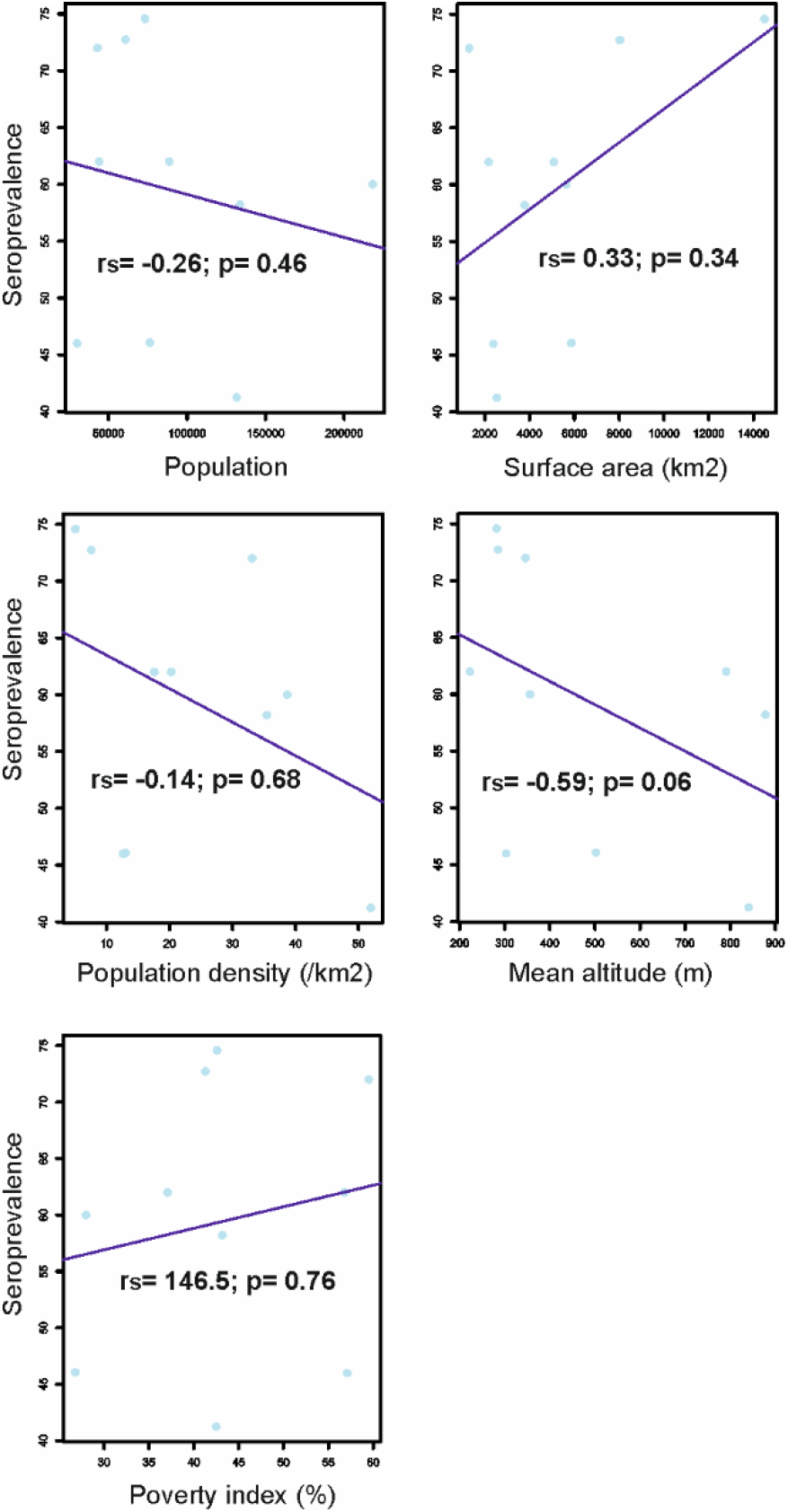
Spearman’s rank correlation test of seroprevalence and population, population density, surface area, poverty index and altitude. Rs= Spearman’s correlation coefficient. Epidemiological data taken from: http://sial.minam.gob.pe/sanmartin/indicador/850, https://www.citypopulation.de/en/peru/sanmartin/admin/

